# Transcriptomics of Bronchoalveolar Lavage Cells Identifies New Molecular Endotypes of Sarcoidosis

**DOI:** 10.1101/2020.07.24.20161448

**Authors:** Milica Vukmirovic, Xiting Yan, Kevin F. Gibson, Mridu Gulati, Jonas C. Schupp, Giuseppe DeIuliis, Taylor S. Adams, Buqu Hu, Antun Mihaljinec, Tony Woolard, Heather Lynn, Nkiruka Emeagwali, Erica L. Herzog, Edward S. Chen, Alison Morris, Joseph K. Leader, Yingze Zhang, Joe G. N. Garcia, Lisa A. Maier, Ron Colman, Wonder P. Drake, Michael Becich, Harrison Hochheiser, Steven R. Wisniewski, Panayiotis V. Benos, David R. Moller, Antje Prasse, Laura L. Koth, Naftali Kaminski

## Abstract

Sarcoidosis is a multisystem granulomatous disease of unknown origin with a variable and often unpredictable course and pattern of organ involvement. In this study we sought to identify specific bronchoalveolar lavage (BAL) cell gene expression patterns indicative of distinct disease phenotypic traits.

RNA sequencing by Ion Torrent Proton was performed on BAL cells obtained from 215 well characterized patients with pulmonary sarcoidosis enrolled in the multicenter Genomic Research in Alpha-1 Antitrypsin Deficiency and Sarcoidosis (GRADS) study. Weighted Gene Co-expression Network Analysis (WGCNA) and non-parametric statistics were used to analyze genome wide BAL transcriptome. Validation of results was performed using a microarray expression data set of an independent sarcoidosis cohort (Freiburg, Germany (n=50)).

Our supervised analysis found associations between distinct transcriptional programs and major pulmonary phenotypic manifestations of sarcoidosis including; TH1 and TH17 pathways associated with hilar lymphadenopathy; TGFB1 and MTOR signaling with parenchymal involvement, and IL7 and IL2 with airway involvement. Our unsupervised analysis revealed gene modules that uncovered four potential sarcoidosis endotypes including hilar lymphadenopathy with increased acute T cell immune response; extraocular organ involvement with PI3K activation pathways; chronic and multiorgan disease with increased immune response pathways; and multiorgan with increased IL-1 and IL-18 immune and inflammatory responses. We validated the occurrence of these endotypes using gene expression, pulmonary function tests and cell differentials from Freiburg. Taken together our results identify BAL gene expression programs that characterize major pulmonary sarcoidosis phenotypes and suggest the presence of distinct disease molecular endotypes.

**Take home message:** Genome wide BAL transcriptomics identified novel gene expression profiles associated with distinct phenotypic traits in sarcoidosis and is suggestive of the presence of novel molecular and clinical sarcoidosis endotypes that could help with further understanding of this heterogenous disease.

## INTRODUCTION

Sarcoidosis is a granulomatous disease of unknown etiology that can affect almost every organ, but affects the lungs in majority of the cases (>90%). The patterns of organ involvement and disease course are often unpredictable, but a substantial number of patients suffer either a relapsing or progressive course with mortality estimated at 12% in advanced cases [1–3]. The etiology of sarcoidosis is still unknown. Despite significance advances in understanding the contribution of genetic predisposition, immune aberrations and the presence of microbial antigens in patients with sarcoidosis, little is known about the genetic networks and environmental factors that determine the phenotype in sarcoidosis. Similarly, treatment is still based on immunosuppression, with corticosteroids serving as first line, and then use of ‘steroid sparing’ agents with limited evidence of long term benefit [4, 5]. Genetics and genomics studies on sarcoidosis have focused on identifying DNA variants or gene signatures associated with sarcoidosis [6–12]. Genome-wide association studies (GWAS) have found associations between MHC region and HLA-DRB1 variants and disease severity and sarcoidosis risk. Most previous whole transcriptome studies focused on identifying gene signatures that distinguish sarcoidosis from control; progressive from non-progressive disease; or from other granulomatous diseases such as tuberculosis [6–13]. While highly informative, they were mostly focused on the peripheral blood, limited in size, heterogeneity of disease manifestations and depth of phenotyping.

In this study, we performed a genome-wide transcriptome analysis of bronchoalveolar lavage (BAL) samples collected from a large cohort of well characterized sarcoidosis patients recruited by the Genomic Research in Alpha-1 Antitrypsin Deficiency and Sarcoidosis (GRADS) study [14]. We conducted both supervised and unsupervised analysis on the measured gene expression profiles of these BAL samples to identify gene signatures that are associated with the heterogeneity in the clinical and phenotypic manifestations of sarcoidosis. The results suggest the presence of novel molecular and clinical endotypes of sarcoidosis.

## METHODS

### GRADS Patient Population

BAL samples were available from subjects enrolled in the GRADS study as previously described [14]. After informed consent and recruitment screening (Supplementary Figure S1) patients were grouped into predefined phenotypic groups based on a modified organ assessment instrument developed by the ACCESS study [15] and described by us in detail [14]. The collected phenotypic information according to the GRADS study protocol include physiological parameters, high-resolution computed tomography (HRCT), BAL, and detailed questionnaires assessing dyspnea, fatigue and quality of life, current and past medical information, occupational, and environmental exposures among others. All recruitment and consenting procedures were compliant with current HIPAA regulations and approved by the local institutional review board (IRB) [14].

### Freiburg Validation Cohort

BAL gene expression data from Affymetrix Human Gene 1.0 ST Arrays were available from 50 individuals with Sarcoidosis recruited independently in Freiburg, Germany. The consents were collected following institutional IRB protocols.

### Sample Preparation and Transcriptomic Analysis

Total RNA was extracted as previously described [16] (Supplement Material). For GRADS samples cDNA libraries were generated, amplified and sequenced on the Ion Proton™ System for Next-Generation Sequencing to obtain sequencing depth of ∼30 million single-end reads/sample with an average read length of 150bps (Supplementary Table S1). Gene expression in Freiburg samples was quantified using the Affymetrix Human Gene 1.0 ST Arrays (Supplementary Material).

### Statistical Analysis

The supervised analysis identified gene signatures associated with predefined phenotypical traits using non-parametric tests (Supplementary Material). Unless otherwise stated in the text we used the false discovery rate (FDR) to control for multiple testing error. The unsupervised analysis used the weighted gene co-expression network analysis (WGCNA)[17]. Genes from chosen WGCNA modules (p<0.05) were used to cluster the patients into subgroups using K-means clustering (Supplementary Material). Chi-square test and Wilcoxon rank sum test were used to assess the significance for categorical and continuous patient characteristics (p<0.05), respectively, amongst the clusters for chosen gene modules (Supplementary Material). The MetaCore™ of GeneGO, Inc. was applied to identify significant enriched pathways (FDR<0.05) for each gene module identified by the unsupervised analysis. The data are publicly available on the GEO database (http://www.ncbi.nlm.nih.gov/geo/) under the accession number GSE109516. All analysis codes and results are available on github page (https://yale-p2med.github.io/SARC_BAL).

## RESULTS

### Patient Cohorts

GRADS cohort: Based on RNA quality and quantity as well as the quality of RNA-Seq data of BAL samples, we included 209 BAL samples from 8 predefined clinical phenotype groups [14]: 25 stage I, 34 stage II-III treated, 42 stage II-III untreated, 19 stage IV treated, 12 stage IV untreated, 14 acute sarcoidosis, 40 remitting and 23 subjects with multiorgan involvement (Table S1). 53.6% were female and 23.4% were black (Table 1a). For the Freiburg cohort, 50 BAL samples were available, 100% of patient were white, 32 were male and 18 were female. The clinical information collected included demographics and 12 clinical phenotypic traits overlapping with the GRADS protocol (Table 1b).

**Table 1a.** Demographic and clinical characteristics of the GRADS cohort categorized by the phenotypic groups.

| PHENOTYPE GROUPS | 1. MULTIORGAN | 2. NON ACUTE<br>STAGE I<br>UNTREATED | 3. STAGE II-III<br>TREATED | 4. STAGE II-III<br>UNTREATED | 5. STAGE IV<br>TREATED | 6. STAGE IV<br>UNTREATED | 7. ACUTE<br>UNTREATED | 8.<br>REMITTING<br>UNTREATED |
| --- | --- | --- | --- | --- | --- | --- | --- | --- |
| **TOTAL, n** | 23 | 25 | 34 | 42 | 19 | 12 | 14 | 40 |
| **SCADDING** |  |  |  |  |  |  |  |  |
| Stage 0, n (%) | 4 (17.4%) | 0 (0.0%) | 1 (2.9%) | 0 (0.0%) | 0 (0.0%) | 0 (0.0%) | 2 (14.3%) | 14 (35.0%) |
| Stage 1, n (%) | 7 (30.4%) | 24 (96.0%) | 0 (0.0%) | 0 (0.0%) | 0 (0.0%) | 0 (0.0%) | 9 (64.3%) | 14 (35.0%) |
| Stage 2, n (%) | 6 (26.1%) | 0 (0.0%) | 23 (67.7%) | 34 (81.0%) | 0 (0.0%) | 0 (0.0%) | 3 (21.4%) | 7(17.5%) |
| Stage 3, n (%) | 4 (17.4%) | 0 (0.0%) | 10 (29.4%) | 8 (19.0%) | 0 (0.0%) | 0 (0.0%) | 0 (0.0%) | 5 (12.5%) |
| Stage 4, n (%) | 2 (8.7%) | 0 (0.0%) | 0 (0.0%) | 0 (0.0%) | 19 (100%) | 12 (100%) | 0 (0.0%) | 0 (0.0%) |
| NA, n (%) | 0 (0.0%) | 1 (4.0%) | 0 (0.0%) | 0 (0.0%) | 0 (0.0%) | 0 (0.0%) | 0 (0.0%) | 0 (0.0%) |
| **FEMALE GENDER, n (%)** | 12(52.2%) | 18(72.0%) | 18(52.9%) | 19(45.2%) | 9(47.4%) | 6(50.0%) | 9(64.3%) | 21(52.5%) |
| **AGE, years (mean ± SD)** | 54.6±10.8 | 46.2±10.9 | 54.3±9.9 | 50.2±9.3 | 52.9±8.0 | 49.8±10.4 | 51.4±12.2 | 52.2±9.8 |
| **AGE, range, years** | 30.9-69.6 | 27.3-64.9 | 34.8-74.9 | 26.0-68.2 | 39.3-69.5 | 32.7-63.6 | 30.8-66.9 | 26.3-71.5 |
| **RACE** |  |  |  |  |  |  |  |  |
| Caucasian/White, n | 15 (65.2%) | 21 (84.0%) | 22 (64.7%) | 31 (73.8%) | 13 (68.4%) | 9 (75.0%) | 9 (64.3%) | 33 (82.5%) |
| African American/Black, n | 8 (34.8%) | 4 (16.0%) | 8 (23.5%) | 10 (23.8%) | 6 (31.6%) | 3 (25.0%) | 4 (28.6%) | 6 (15.0%) |
| Other, n | 0 (0.0%) | 0 (0.0%) | 4 (11.8%) | 1 (2.4%) | 0 (0.0%) | 0 (0.0%) | 1 (7.1%) | 1 (2.5%) |
| **FEV1 (L), mean ± SD** | 2.6±1.3 | 2.8±0.8 | 2.3±0.8 | 2.9±0.8 | 2.08±0.8 | 2.4±1.9 | 3.0±0.6 | 3.1±0.9 |
| **FEV1% PRED, mean ± SD** | 90.0±13.4 | 84.5±30.9 | 78.6±20.2 | 84.24±26.2 | 69.5±21.3 | 84.2±25.9 | 86.1±39.4 | 96.2±21.4 |
| **FVC (L), mean ± SD** | 3.4±1.6 | 3.7±1.0 | 3.2±1.1 | 3.9±1.0 | 3.1±0.9 | 3.5±2.4 | 3.7±0.8 | 4.0±1.2 |
| **FVC% PRED, mean ± SD** | 89.2±11.4 | 84.8±29.4 | 82.5±18.8 | 85.5±24.4 | 78.2±16.2 | 91.3±23.9 | 83.1±38.0 | 93.9±20.4 |
| **DLCO(ml/min/mmHg), mean±SD** | 23.5±8.6 | 21.8±10.6 | 20.1±9.5 | 23.8±8.8 | 21.7±8.9 | 20.7±11.8 | 19.5±11.0 | 27.3±9.9 |
| **DLCO%, mean±SD** | 86.1±24.1 | 85.8±26.4 | 69.5±28.1 | 81.0±24.5 | 72.4±18.1 | 67.3±28.1 | 74.1±37.3 | 90.7±23.9 |

*Definition of abbreviations:* PFT% PRED: pre-bronchodilator PFT in predicted percentage.

**Table 1b.** Patient characteristics of the GRADS and Freiburg cohort for the 12 overlapping clinical features.

|  | GRADS | Freiburg | P value |
| --- | --- | --- | --- |
| TOTAL, n | 209 | 50 |  |
| **SCADDING** |  |  | 0.02 |
| Stage 0, n (%) | 21 (10.1%) | 0 (0.0%) |  |
| Stage 1, n (%) | 54 (25.8%) | 5 (10.0%) |  |
| Stage 2, n (%) | 73 (34.9%) | 28 (56.0%) |  |
| Stage 3, n (%) | 27 (12.9%) | 11 (22.0%) |  |
| Stage 4, n (%) | 33 (15.8%) | 6 (12.0%) |  |
| NA, n (%) | 1 (0.5%) | 0 (0.0%) |  |
| **FEMALE GENDER, n (%)** | 112 (53.6%) | 18 (36.0%) | 0.04 |
| **AGE, years (mean ± SD)** | 51.6±10.2 | 46.3±13.2 | 2.9x10<sup>-3</sup> |
| **AGE, range** | 26.0-74.9 | 21.0-81.0 |  |
| **FEV1 (L), mean ± SD** | 2.7±1.0 | 2.6±0.9 | 0.497 |
| **FEV1% PRED, mean ± SD** | 85.1±25.3 | 72.9±20.9 | 2.0x10<sup>-4</sup> |
| **FVC (L), mean ± SD** | 3.6±1.2 | 3.5±1.1 | 0.45 |
| **FVC% PRED, mean ± SD** | 86.5±23.0 | 81.4±20.3 | 0.05 |
| **FEV1/FVC ratio, mean ± SD** | 0.8±0.1 | 0.8±0.1 | 0.92 |
| **MACROPHAGE %, mean±SD** | 73.2±32.0 | 54.2±21.2 | 2.6x10<sup>-9</sup> |
| **LYMPHOCYTE, mean¬±SD** | 10.5±11.5 | 41.5±21.6 | 1.4x10<sup>-19</sup> |
| **NEUTROPHIL, mean¬±SD** | 1.2±2.5 | 2.8±4.4 | 9.0x10<sup>-5</sup> |
| **EOSINOPHIL, mean¬±SD** | 0.21±0.95 | 1.08±1.29 | 6.1x10<sup>-13</sup> |

### Gene expression patterns associated with hilar lymphadenopathy, parenchymal or airway involvement

We assessed the association of gene expression of 209 BAL samples with 24 predefined clinical traits including Scadding staging, PFTs, CT parameters, age, gender, BAL cell counts and the eight phenotypic groups. This analysis revealed distinct genes associated with most traits (FDR<0.05) as well as overlapping genes (Figure 2a, Supplementary tables).

**Figure 1.**
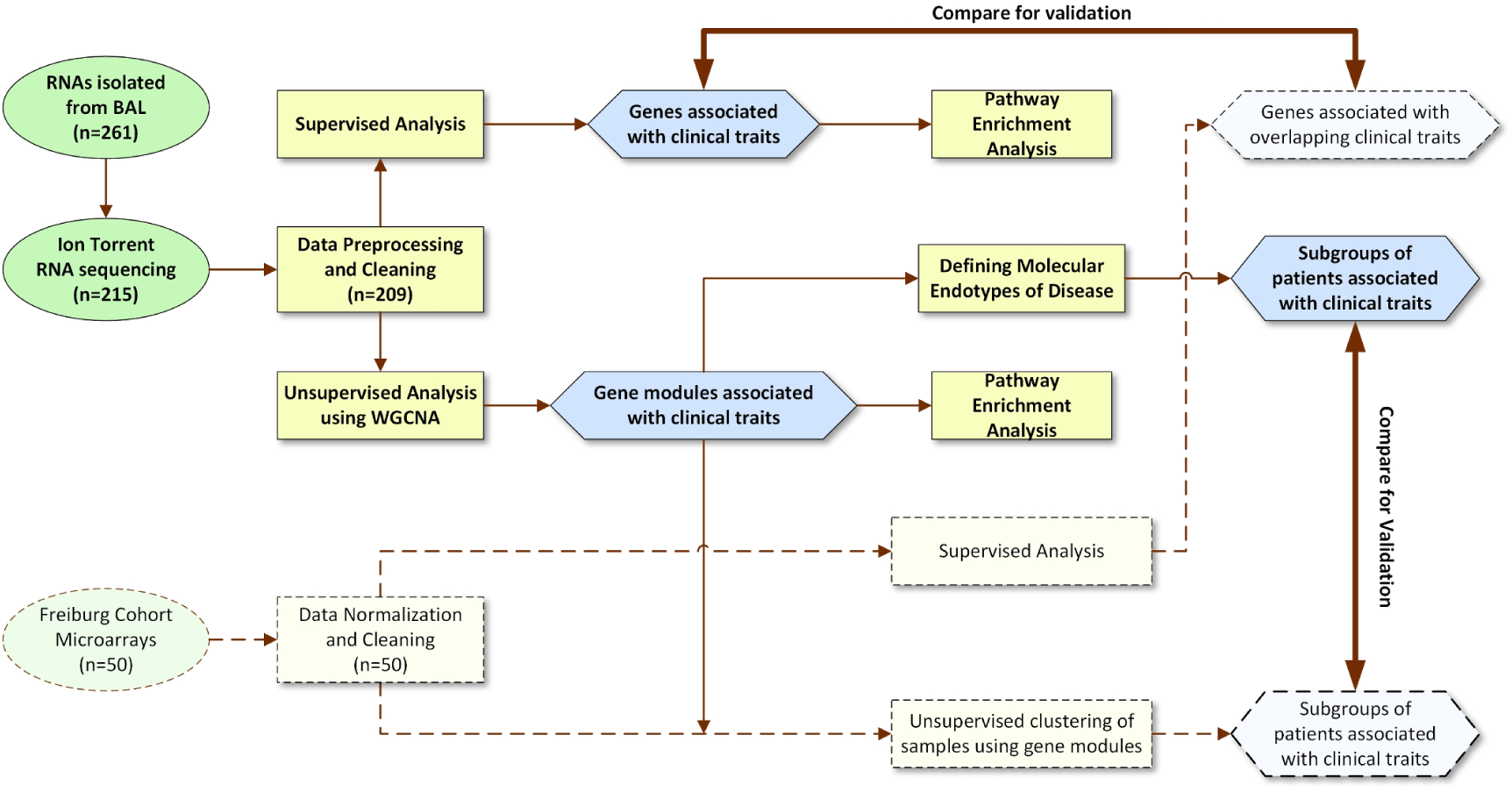
Study workflow.

**Figure 2.**
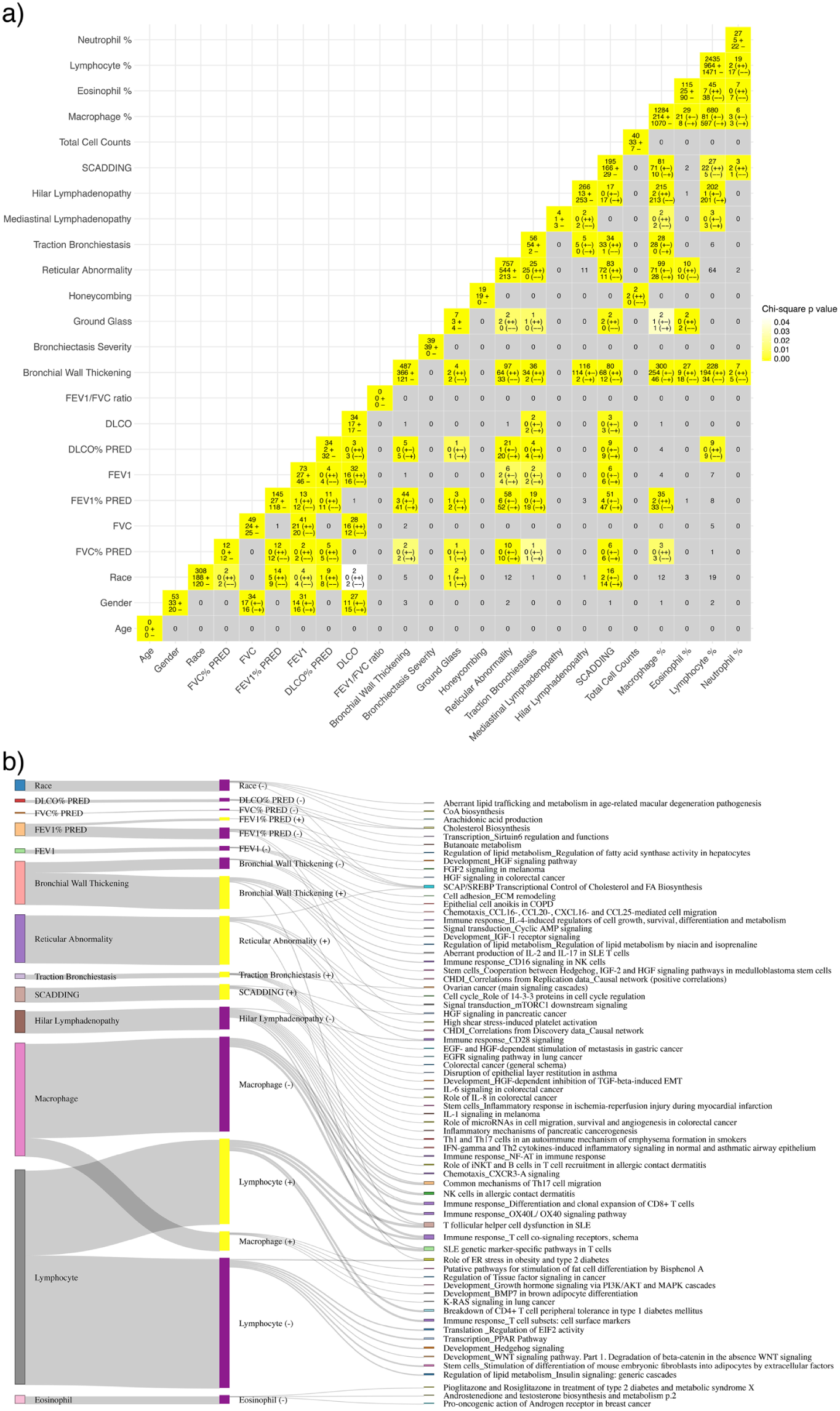
Association of gene expression and clinical variables in supervised analyses. a) The number of genes shared between any two traits from a given set of clinical traits are shown in a squared matrix format with columns and rows both representing clinical traits. Entries colored in grey have zero overlap or FDR<0.05. The color of entries with non-grey color (white to yellow) describes the significance of overlap by Chi-suqare test. Each entry describes the number of genes in total in the first fow followed by rows describing the number of genes with a given direction of correlation in the parenteses (positive and negative correlations depicted by (+) and (-), respectively). b) The top 5 significant (FDR<0.05) enriched pathways for genes associated with each clinical trait identified by GeneGo Metacore (Thomson Reuters) are shown as a Sankey plot. Genes significantly associated with each clinical trait are represented by bars on the left. These genes were further divided into positively and negatively correlated genes represented by bars in the middle colored in yellow and purple respectively. Each set of negatively or positively correlated genes was further connected to its signifticantly (FDR<0.05) enriched pathways (bars on the right). The lengths of all bars are proportional to the number of genes represented. Only the top 5 significant (FDR<0.05) pathways with at least 3 genes correlated are shown.

Increased Scadding stage, was significantly associated with increased expression in 166 genes and decreased expression in 29 genes (Figure 2a). Of these genes, many were also significantly associated with CT traits such as hilar lymphadenopathy (17 genes), and reticular abnormalities (83 genes). In the presence of both progressive Scadding staging and more reticular abnormality, increased genes include PLA2G7, ID1, LGMN, CCL2 and decreased genes include PDLIM1 and AOC3. SLC40A1 was increased with progressive Scadding staging, more reticular abnormality and more hilar lymphadenopathy. These genes are known to be involved in fibrosis and chronic obstructive lung disease [18–20]. Genes increased with progressive Scadding staging were enriched for IL1, IL8 and IL6 pathways (Figure 2A), previously associated with inflammatory bowel disease [21] and lung fibrosis [18]. Interestingly, many of these genes are also increased with changes in clinical traits not included in the definition of Scadding stage, such as increased bronchial wall thickening (68 genes; TREM2, CHIT1, LGMN) and decreased FEV1% and FVC% (47 and 6 genes; IDI1 and INSIG1), previously reported in sarcoidosis [22, 23].

Among the CT phenotypic traits, reticular abnormality, hilar lymphadenopathy, and bronchial wall thickening were significantly associated with large numbers of genes (757, 266 and 487 respectively, Figure 2a). Genes increased in the presence of hilar lymphadenopathy were enriched for Th1 and Th17, IFN-gamma, and NFAT signaling (Figure 2b). These include genes CD28, STAT1, CXCR3, CCR4 known to drive T cell receptor and Interferon signaling as well as the TH1/TH2 balance and previously reported in sarcoidosis [24, 25]. Genes increased with more severe bronchial wall thickening (Figure 2b) are enriched for aberrant IL-2 and IL-7 pathways including MRC2, SLC40A1, F2R, IL7, PTPN7, ADORA2A, SPRY2, PLA2G7, PTGS1. Reticular abnormalities on CT scan were positively correlated with many known fibrosis associated genes such as TGFBR1, COL3A1, TLR3, ID1, TCF4, IGFBP6, PLA2G7, FADS1, ARGHAP12, and MMP10 [26–29]. Interestingly, ID1, HMGC1, SEPP1 were also inversely correlated with DLCO% and FVC%, potentially reflecting the shared biology underlying physiological restriction, loss of diffusion capacity and reticular abnormalities. Pathway analysis of positively correlated genes with reticular abnormality revealed an enrichment for MTOR (Cytochrome C, SC5D, HIF1A, PPAR alpha), and cell cycle (CDC25C, CHK2, CDK1) signaling that was also seen in genes increased with progressive Scadding staging (Figure 2b).

Taken together, our results suggest that different transcriptional programs affect the three major phenotypic manifestations of pulmonary involvement in sarcoidosis with TH1, TH17 associated with hilar lymphadenopathy, TGFB1 and MTOR signaling for parenchymal involvement and IL7 and IL2 for airway involvement.

### Genes associated with race are also associated with hilar lymphadenopathy, parenchymal or airway involvement

308 genes were associated with race (188 increased in white and 120 in black; Figure 2A). SLC22A16, NME4, PWP2, ASRGL1 and SCARB1 were the top increased genes in blacks while CD300C, LAMA1, RNF135, ST14 were the top genes increased in whites (Figure 2a). Expression of SLC22A16 and PWP2 were previously shown to be race dependent in cancer patients [30]. Aberrant lipid trafficking, CoA biosynthesis and arachidonic acid production were enriched in the genes higher in black while no enriched pathway was found for the genes higher in white. Genes increased in blacks had a significant overlap with genes increased with progressive Scadding stage (MYOZ1, SQRDL, FAM213A), decreased PFTs (TCEA3, FAM213A, MYO1E, CYP51A1, SQLE) being consistent with findings from previous studies on the importance of race in the disease severity [31]. Genes increased with reduced lung function (DLCO%, FVC%, FEV1%), increase in reticular abnormality and in black subjects were associated with SCAP/SREBP transcriptional control of cholesterol and fatty acid biosynthesis (Figure 2b), potentially reflecting race specific pathways of injury.

### Genes increasing with higher lymphocyte fraction in BAL reveal shared transcriptional programs related to hilar lymphadenopathy and bronchial wall thickening

Disease activity is known to be associated with changes in BAL cell composition [32]. While total BAL cell count was associated with only a small number of genes, lymphocytes and macrophages fractions in BAL were associated with the largest number of associated genes (2,435 and 1,284 respectively, Figure 2a, Supplement Tables). Among the top genes (Spearman’s rho > 0.3, FDR < 0.05) positively correlated with increased lymphocyte count were known markers of T lymphocyte subpopulations such as CD2, CD3, CD6, CD5, CD96 and CD247 [33–36]. Amongst other positively correlated genes were: markers of lymphocyte activation (ITK, LCK, CD28, CTLA4, IL2RB), the granzymes GZMA, GZMB and GZMH, ETS1, the chemokines and their receptors (CCL5, CXCL9, CCR4, CXCR3,4,6) and cytokines such as IFNG, IL6, IL26, IL32 and their receptors (IL2RB,G, IL12RB2, IL15RA, IL18RA, IL21RA), and inflammatory regulators (JAK3, NFATC and NKB2). Genes significantly correlated with lymphocyte count significantly overlapped with those genes associated with bronchial wall thickening, hilar lymphadenopathy, Scadding but not with genes associated with reticular abnormality, PFT and demographics (Figure 2A). Among the genes positively correlated with both lymphocyte count and bronchial thickening were CCR5, CCR6, CD84, CD28, IL12RB, IL18R, IL21R and IFNG, potentially reflecting activation of CD4+ T lymphocytes, T:B lymphocytes interactions, Th1 inflammatory response and susceptibility to sarcoidosis [37–40]. The increased lymphocyte count associated genes that overlapped with increased hilar lymphadenopathy were LY9, GNAO1, IFNG, F2R, CCR 4/5/8), CD6, CD5 and KIF21B [41]. Genes associated with higher lymphocyte fractions were associated with numerous immune T cell responses (TH1/TH17, INF-gamma, OX40L/OX40), follicular T-Helper cell dysfunction, T-cell co-signaling receptors, and SLE genetic marker genes (Figure 2b, Supplement Tables).

### WGCNA gene modules associate significantly with PFTs, CT imaging features and BAL cell differentials

The WGCNA analysis identified 48 gene modules and evaluated the correlation of their eigen genes with PFTs, CT imaging features (mediastinal and hilar lymphadenopathy, traction bronchiectasis, micronodule, ground glass, reticular abnormality) and BAL cell differentials (Supplementary Figure E3). Among the 48 modules, 5 modules (module 1, 4, 18, 33 and 47) were significantly correlated (p value<0.05) with multiple clinical traits and we focused on them for further investigation (Figure 3a).

**Figure 3.**
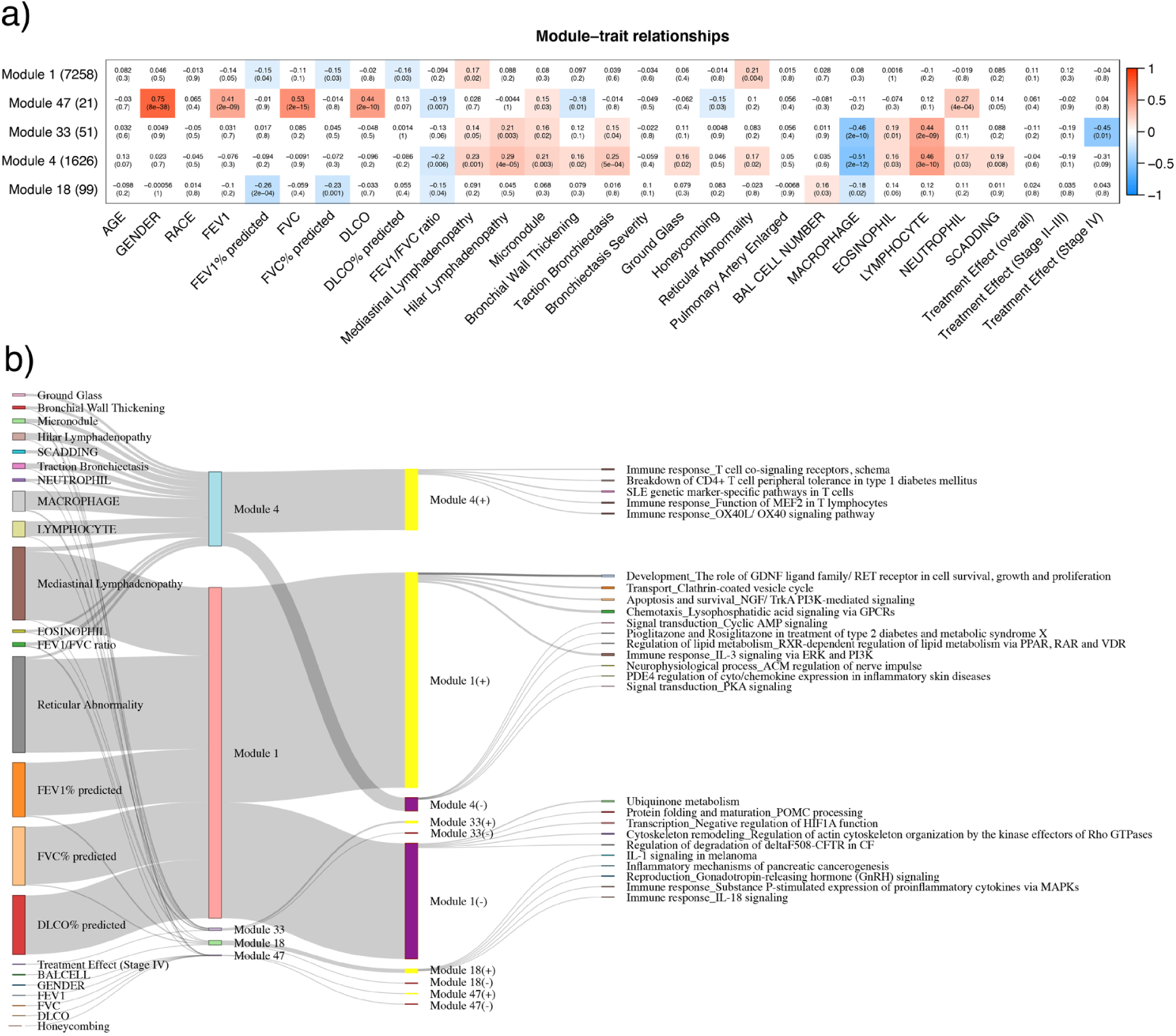
Identification of novel endotypes of sarcoidosis using WGCNA. a) Heatmap showing the correlation between the 5 chosen gene modules and the clinical traits, including demographics, PFTs, CT imaging measures, BAL cell differentials, Scadding stage and treatment within stage III and IV. The correlation coefficient value and the corresponding p value (in parentheses) are shown in each entry. b) Sankey plot visualizing the correlation between gene modules and clinical traits as well as the significant enriched pathways for each gene module. Only the top 5 significant (FDR<0.05) pathways with at least 3 overlapping genes are shown. The positive and negative signs in parentheses represent positively and negatively correlated genes for continuous phenotypes, or increased and decreased for binary phenotypes, respectively. Details of the gene modules can be found at https://yale-p2med.github.io/SARC_BAL.

Module 4 (1,626 genes) was positively correlated with most CT imaging features, BAL cell differentials and Scadding staging suggesting a plausible link between BAL gene expression, increased Scadding stage, increased lymphocytes and eosinophils differentials, and negatively correlated with macrophage differential and FEV1/FVC ratio (Figure 3a). Module 1 had the largest number of genes (7,258) was negatively correlated with all PFT% predicted, and positively correlated with the presence of mediastinal lymphadenopathy and reticular abnormality. No correlation was found for BAL cell differentials or the total BAL cell counts suggesting the existence of large number of gene signatures associated with lung function regardless of the BAL cell differentials. In addition, module 18 (99 genes) was negatively correlated with FEV1% predicted, FVC% predicted and FEV1/FVC ratio as well as macrophage differential. Module 33 (51 genes) was positively correlated with the presence of mediastinal and hilar lymphadenopathy, micronodule, traction bronchiectasis, lymphocyte and eosinophil cell differentials (Figure 3a). Module 47 contained 21 genes that correlated with sex or with absolute PFT but not % predicted PFT, reflecting the effect on sex on PFT.

### Four gene modules are suggestive of novel molecular endotypes of sarcoidosis

To define novel disease endotypes we performed K-means clustering analysis using genes from each of the WGCNA modules to identify clusters of sarcoidosis patients with distinct clinical traits (Figure 4).

**Figure 4.**
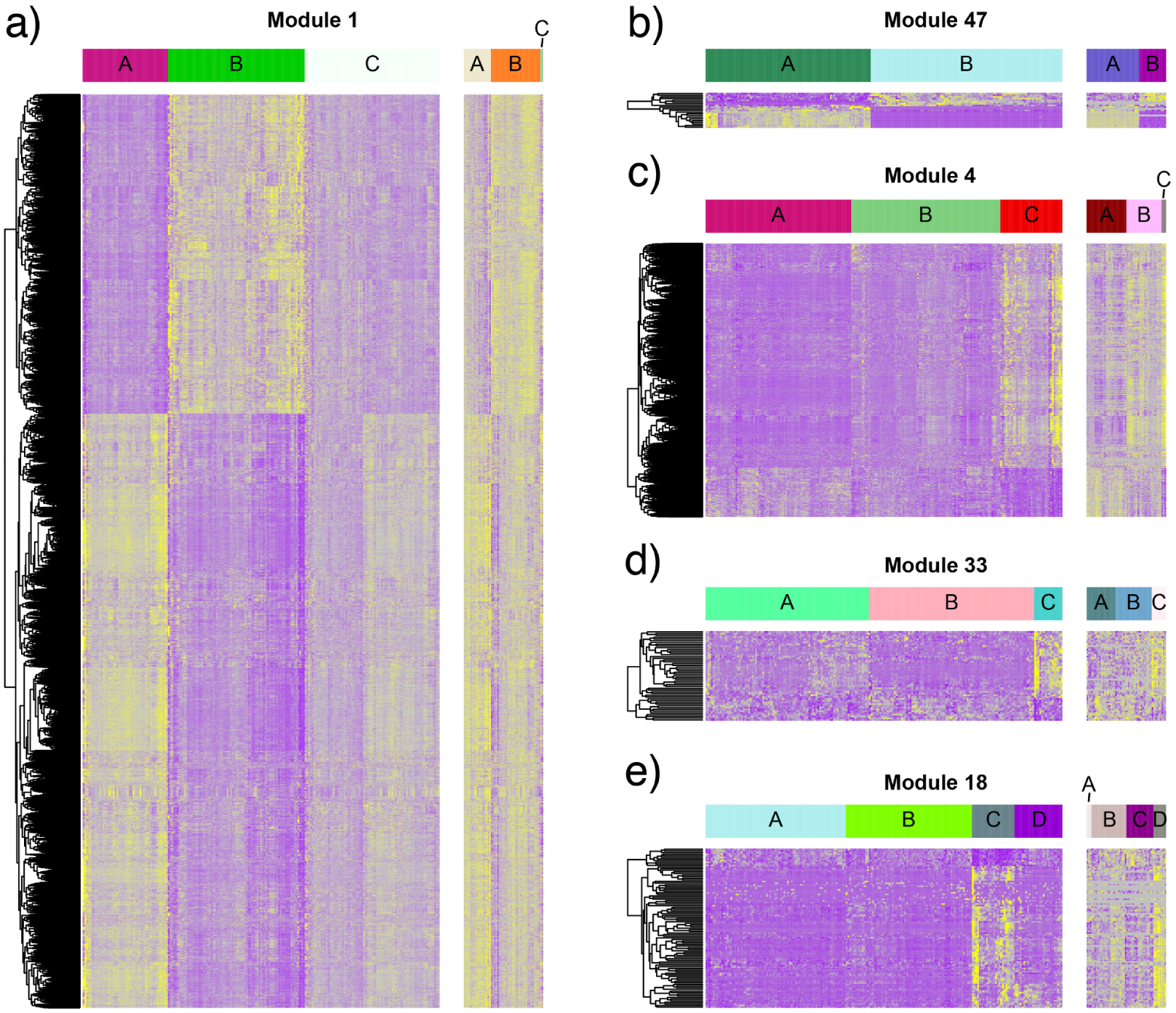
Heatmaps showing the expression pattern of the chosen gene modules and their corresponding K-means clustering results on the patients from GRADS cohort and Freiburg cohort. Panels a) to e) are heatmaps of the module 1, 47, 4, 33, and 18, respectively. The patient clusters identified by K-means are labelled as cluster A, B, C and D on the top of the heatmaps. In each panel, the left heatmap is for GRADS cohort and the one on the right is for the Freiburg cohort. Genes in these two heatmaps are shown in the same order.

Clustering based on the genes in Module 4 identified clusters of patients who differed clinically with one group having significantly more hilar and mediastinal lymphadenopathy, larger lymph nodes, increased Scadding stage, less remitting phenotype and more lymphocytes in BAL (Figure 4c, cluster C) than the others. These patients had a history of increased exposure to woodfire smoke and had more skin and kidney involvement. Increased T cell immune response and decreased signal transduction via cAMP and PKA was observed in this cluster of patients suggesting that these patients have an acute lymphocytic inflammation (Figure 4c, Supplementary Table E3).

Clustering patients based on genes in module 1 revealed clusters of patients who differed clinically and environmentally with one group having significantly more mediastinal lymphadenopathy and reticular abnormality in the lung, less multi-organ phenotype (affected eyes), more environment effects and with more subjects living in Arizona and Tennessee. Up-regulation of apoptotic, immune response and development pathways related to PI3K activation was observed in the same cluster of patients (Figure 4a, cluster A, Supplementary Table E3).

Clustering patients based on genes in module 33 revealed clusters of patients who differed clinically with one group, cluster C, having significantly more lymphocytes in BAL and more lymph organ involvement, more exposure to sand, less fatigue and did less work in the house than patients in cluster B. Patients in cluster C were more chronic and had higher multiorgan involvement than cluster A (Figure 4d, Supplementary Table E3).

Clustering patients based on genes in module 18 identified clusters of patients who differed clinically and environmentally with one distinguished group (Figure 4e, cluster C) having significantly more patients from Connecticut, New Jersey, New York, Pennsylvania and Tennessee, more lung, joint and kidney involvement, increased urinary calcium and were exposed to more wood coal before diagnosis. However, these patients have less mediastinal lymphadenopathy and micronodules than patients in cluster B (Figure 4e, Supplementary Table E3). Increased IL-1 and IL-18 immune and inflammatory responses were observed in cluster C (Figure 3b).

### Novel sarcoidosis endotypes validated using an independent cohort

To validate the endotypes we discovered, we used the genome wide BAL transcriptome data from an independent cohort of sarcoidosis patients (Freiburg cohort). The Freiburg patients were clustered using genes from each of the 4 novel modules and gender module independently (Figure 4). Available Scadding stage, age, gender, BAL cell differential and PFTs were compared between the two extreme patient clusters defined by each module in both GRADS and Freiburg cohorts (Table 2). The comparison showed that the endotype of gender including PFTs (module 47) was fully validated. For the endotype of hilar lymphadenopathy and acute lymphocytic inflammation (module 4), the association with macrophages, lymphocytes, neutrophils, PFTs and age were validated. Module 18, a chronic sarcoidosis endotype, and Module 33, a multiorgan involvement, were validated for age, Scadding, gender, PFTs, macrophages and lymphocytes counts. Module 1, an extraocular organ involvement and PI3K activation was validated for all clinical traits besides FVC% and FEV1%.

**Table 2.** Association between chosen molecular endotypes and the 12 overlapping clinical traits in GRADS and Freiburg cohorts for validation. Each entry is a p value for the difference of the given clinical trait between the two extreme clusters of patients identified using the the corresponding gene module in the corresponding cohort.

|  | Module 47<br>Gender module |  | Module 4<br>Hilar<br>Lymphadenopathy<br>and Acute<br>Lymphocytic<br>Inflammation |  | Module 33<br>Multiorgan<br>involvement with<br>increased immune<br>response |  | Module 18<br>Chronic<br>sarcoidosis |  | Module 1<br>Extraocular organ<br>involvement and<br>PI3K activation |  |
| --- | --- | --- | --- | --- | --- | --- | --- | --- | --- | --- |
|  | GRADS<br>(A vs B)<br>P value | Freiburg<br>(A vs B)<br>P value | GRADS<br>(A vs C)<br>P value | Freiburg<br>(A vs C)<br>P value | GRADS<br>(B vs C)<br>P value | Freiburg<br>(B vs C)<br>P value | GRADS<br>(B vs C)<br>P value | Freiburg<br>(A vs D)<br>P value | GRADS<br>(A vs B)<br>P value | Freiburg<br>(B vs C)<br>P value |
| SCADDING | **0.02** | 0.11 | **0.01** | 0.34 | 0.53 | 0.06 | 0.24 | 0.15 | 0.44 | 0.79 |
| AGE | 0.80 | 0.59 | 0.07 | 0.66 | 0.91 | 0.28 | 0.66 | 0.78 | 0.45 | 0.29 |
| GENDER | **<5x10<sup>-4</sup>** | **2.1x10<sup>-11</sup>** | 0.24 | **9x10<sup>-3</sup>** | 0.43 | 0.29 | 0.64 | 0.08 | 0.86 | 1 |
| MACROPHAGE | 0.75 | 0.05 | **7x10<sup>-8</sup>** | **5x10<sup>-3</sup>** | **2x10<sup>-5</sup>** | **1x10<sup>-3</sup>** | **0.04** | **0.01** | 0.53 | 0.33 |
| LYMPHOCYTES | 0.53 | 0.05 | **9x10<sup>-8</sup>** | **5x10<sup>-3</sup>** | **2x10<sup>-5</sup>** | **1x10<sup>-3</sup>** | **0.03** | **0.01** | 0.35 | 0.44 |
| NEUTROPHILS | 0.16 | 0.47 | **0.01** | **0.03** | 0.06 | **8x10<sup>-3</sup>** | 0.16 | 0.05 | 0.46 | **0.004** |
| EOSINOPHILS | 0.80 | 0.84 | **8x10<sup>-5</sup>** | 0.39 | **0.03** | 0.38 | 0.85 | 0.14 | 0.11 | 0.40 |
| FVC | **9x10<sup>-15</sup>** | **3x10<sup>-3</sup>** | 0.23 | 0.33 | 0.23 | 0.30 | 0.82 | 0.5 | 0.11 | 0.19 |
| FEV1 | **2x10<sup>-9</sup>** | **8x10<sup>-3</sup>** | 0.62 | 0.30 | 0.47 | 0.39 | 0.80 | 0.38 | 0.06 | 0.22 |
| FVC% predicted | 0.98 | 0.14 | 0.79 | 0.64 | 0.97 | 0.08 | 0.82 | 0.33 | **0.01** | 0.19 |
| FEV1% predicted | 0.78 | 0.75 | 0.84 | 0.91 | 0.55 | 0.08 | 0.65 | 0.22 | **0.02** | 0.28 |
| FEV1/FVC ratio | 0.36 | **0.03** | 0.26 | 0.71 | 0.25 | 0.72 | 0.29 | 1.00 | 0.90 | 0.87 |

We also found overlap in genes significantly associated with FVC% predicted and FEV1% predicted in both cohorts with 16 and 39 genes for FVC% predicted and FEV1% predicted, respectively (chi-square p value=2.22e-16 and 7.41e-12). Amongst the top negatively correlated genes with FVC% in both cohorts were: COL1A2, IGFBP6, MAT2A, AQP9, GJA1 and EPDR1 suggestive of involvement of IGF family signaling pathways. The 39 genes highly negatively correlated with FEV1% predicted included: CYP51A1, FADS1, COL1A1, HSPA7, LDLR, and NFKB1 suggestive of involvement of NF-kB related pathways.

## DISCUSSION

Our study based on genome-wide transcriptome analysis of 209 BAL samples represents the first and the largest effort to examine the BAL transcriptome in sarcoidosis with pulmonary involvement. Using both, supervised and unsupervised analysis, our study revealed specific gene profiles correlated with activity and severity of disease including immune, inflammatory and profibrotic mediators. Most importantly, our study identified four new groups of sarcoidosis patients with specific molecular, clinical and environmental characteristics.

The unsupervised analysis identified 4 gene modules that were strongly correlated with CT imaging features of pulmonary involvement, Scadding stage, and BAL cell differentials. These modules further identified groups of patients with 1) hilar lymphadenopathy and acute lymphocytic inflammation, 2) extraocular organ involvement and PI3K activation, 3) chronic sarcoidosis, 4) multiorgan involvement with increased immune response (Figures 3 and 4). Identification of BAL gene modules represents a robust and novel molecular approach to address clinical heterogeneity of sarcoidosis patients initially classified into 8 clinical phenotypes in GRADS study [14]. BAL gene modules had the strongest correlation with multiple clinical features suggestive of pulmonary involvement which might be the top factor to consider for further patient phenotyping, as previously suggested [42]. Gender response gene module clearly separated female from male patients based on 15 out of 21 gene expression, suggesting on accuracy and sensitivity of our method (Figure 3 and 4 and Supplement table E3). Previously, unsupervised clustering approaches used solely clinical patient characteristics to successfully subgroup sarcoidosis patients based on organ involvement (lung, abdominal, heart, muscle and extrapulmonary) or to modulate sarcoidosis based on personal and environmental factors [43, 44]. Our study is the first to combine clinical and environmental factors with BAL transcriptome data to phenotype sarcoidosis with pulmonary involvement.

Supervised analysis identified numerous divergent and convergent gene expression patterns associated with hilar lymphadenopathy, parenchymal or airway involvement in sarcoidosis (Figure 2). Our results suggest that different transcriptional programs affect the three major phenotypic manifestations of pulmonary involvement in sarcoidosis with TH1 and TH17 associated with hilar lymphadenopathy, TGFB1 and MTOR signaling for parenchymal involvement, and IL7 and IL2 for airway involvement. These responses were amongst identified pathways and also known to be involved in sarcoidosis pathogenesis [6, 8, 45, 46]. An increase in reticular abnormality observed in chest radiology of sarcoidosis patients was associated with increased molecular signaling in BAL such as: growth factor, apoptosis/survival, mTORC1 and immune CD28 signaling (Figure 2). The activation of these signaling pathways is suggestive of granuloma formation and presence of fibrosis in sarcoid lungs [47–49]. Genes associated with race (black) were also associated with hilar lymphadenopathy, aberrant lipids pathways, parenchymal or airway involvement in sarcoidosis. However, our data do not provide more evidence to demonstrate if this difference is race specific or directly related to the presence of sarcoidosis, as described previously [50] or due to differences in disease severity and manifestations. Genes positively correlated with increased lymphocyte fraction in BAL reveal shared transcriptional programs related to hilar lymphadenopathy and bronchial wall thickening. Genes positively correlated with increased macrophage fraction in BAL are associated with increased cell differentiation, and development pathways such as PI3K/AKT, MAPK, and BMP7 signaling suggesting indirectly that these BAL macrophages were in contact with inflammatory sites and granuloma core in lungs (Supplement Material) [51, 52]. Genes positively correlated with decreased eosinophil fraction in BAL were associated with decreased airway thickness and could reflect the level of lung inflammation, as well as on severity and progression of sarcoidosis (Supplement Material).

Our study has several limitations including samples size, heterogeneity of clinical features and confounding effect of BAL cell differentials. Although our study collected a relatively large sample size for transcriptomics analysis, the unpredictable course and presentation of sarcoidosis that leads to heterogeneity of clinical features limited us in identifying more than 4 distinct endotypes in the GRADS sarcoidosis cohort [14, 53, 54]. Another limitation of our study is the impact of the BAL cell differentials on the gene expression data for gene modules 4 (hilar lymphadenopathy and acute lymphocytic inflammation) and 33 (multiorgan involvement with increased immune response) but not for modules related to gender, chronic sarcoidosis and extraocular organ involvement. We were also not able to provide a complete transcriptomics link to treatment [55] and race effects [56].

In summary, our study identified gene profiles associated with major phenotypic manifestations of pulmonary involvement in sarcoidosis, as well as identified 4 novel endotypes. Future analyses of the GRADS cohort of individuals with sarcoidosis will focus on understanding the link between BAL and blood gene expression and their relationship with phenotypes of sarcoidosis.

## Data Availability

The data are publicly available on the GEO database (http://www.ncbi.nlm.nih.gov/geo/) under the accession number GSE109516. All analysis codes and results are available on github page (https://yale-p2med.github.io/SARC_BAL).

## Acknowledgments

We thank all patients who participate in the GRADS study for contributing samples.

## Authors’ Contributions

NK, LLK, DM, KFJ, WD conceived and designed the experiments; KFG, MG, SMN, MB, HH, EH, ESC, AM, JKL, JGNG, SRW, LAM, DRM, KP, WPD conducted patient phenotyping, classification, supervised sample and data collection; MV, TA, TW and JD performed the RNA sequencing experiments; JS and AP collected and generated the microarray data of the Freiburg cohort for validation; XY, MV, NK, BH, AM, YZ, NE, PVB, JS, AP analyzed the data; NK, XY, MV and LLK supervised the analytic plan; MV, XY and NK wrote the manuscript with input from all other authors. All authors have read and approved the manuscript.

